# Changes in General Practice use and costs with COVID-19 and telehealth initiatives

**DOI:** 10.1101/2022.07.11.22277516

**Authors:** Danielle C Butler, Grace Joshy, Kirsty A Douglas, Muhammad-Shahdaat Bin-Sayeed, Jennifer Welsh, Angus Douglas, Rosemary J. Korda, the ANU Telehealth in Primary Care study group

**Affiliations:** National Centre for Epidemiology and Population Health, Research School of Population Health, Australian National University, Canberra, ACT, Australia; Academic Unit of General Practice, Australian National University, Canberra, ACT

**Keywords:** Telehealth, COVID-19, primary care, out-of-pocket costs, linked data

## Abstract

**Background:** In response to the COVID-19 pandemic, general practice (GP) in Australia underwent a rapid transition, including the rollout of population-wide telehealth, with uncertain impacts on GP use and costs.

**Objective:** To describe how use and costs of GP services in Australia changed in 2020—following the pandemic and introduction of telehealth—compared to 2019, and how this varied across population subgroups.

**Method:** Data for ∼19M individuals from Census 2016 were linked to Medicare data for 2019-2020 through the Multi-Agency Data Integration Project. We used regression models to compare age-sex-adjusted GP use and out-of-pocket cost (OPC) over time, overall and by sociodemographic characteristics.

**Results:** The number of people who visited a GP in Q2-Q4 of 2020 decreased by 4% compared to Q2-Q4 of 2019. The mean number of face-to-face GP services per quarter declined, while telehealth services increased, with overall use of GP services in Q4 2020 similar to or higher than Q4 2019. The proportion of total GP services by telehealth stabilised at ∼25% in Q4 2020. However, individuals aged 3-14 or ≥70 years and those with limited English proficiency used fewer GP services in 2020 compared to 2019, with a lower proportion by telehealth. Mean OPC-per-service was lower across all subgroups in 2020 compared to 2019.

**Discussion:** Introduction of widespread telehealth largely maintained use of GP services during the pandemic and minimised OPCs, but not for all population subgroups. This may indicate technological, social or other barriers in these populations, as well as pandemic-related changes in healthcare use.

**HOW THIS FITS IN:** In response to the COVID-19 pandemic, major telehealth initiatives were implemented to ensure access to primary healthcare while minimising disease transmission. Using routinely collected, whole-of-population data from Australia, we show that the introduction of telehealth during the pandemic largely maintained use of GP services while minimising costs. However, compared to pre-pandemic levels, GP use was lower among individuals aged 3-14 or ≥70 years and those not proficient in English, although these groups also saw the greatest reduction in out-of-pocket cost per service. As telehealth initiatives are integrated into standard GP care, it is vital to ensure telehealth is designed and funded to support these groups and the ongoing financial viability of practices.

## INTRODUCTION

During the COVID-19 pandemic, health systems around the world implemented major telehealth initiatives to ensure access to primary healthcare while minimising disease transmission[1, 2]. In March 2020, Australia introduced new telehealth items[3] to fund primary care teleconsultations (by phone and video) for the whole population through Medicare–Australia’s universal health insurance scheme. Little is known of the potential impact on individual use and costs of primary care and whether certain population subgroups were differentially affected.

Prior to the pandemic, funding for telehealth in Australia was limited to certain jurisdictions or select specialist services[4], with telehealth services accounting for less than 1% of Medicare-subsided consultations[5]. However, just two months after the introduction of the new telehealth items, telehealth consultations constituted 36% of all general practice (GP) consultations, stabilising over the remainder of 2020/21 to ∼20% of services[6]. Similar patterns of use have been observed internationally[7-10].

Despite high levels of overall use of telehealth, international data have shown that uptake is not necessarily even across population groups, potentially exacerbating pre-existing inequalities in access to care. In the US, children and adolescents, older people, and low income and ethnically and linguistically diverse groups have lower uptake of telehealth [7, 11, 12]. Data from primary care clinics limited to specific jurisdictions, covering ∼30% of the total Australian population, have shown that older people, males, and people living in socioeconomically disadvantaged areas have lower uptake of telehealth [13]. Few studies have examined changes in healthcare costs since large-scale telehealth introduction [11, 14, 15] and none have investigated whether this varies across population subgroups. Up until October of 2020 Medicare-subsided telehealth services in Australia were required to be provided at no additional cost to the patient. Yet how average costs of GP services changed in relation to this is unknown.

In this study, we examine how individual use of GP services and associated costs changed in the latter part of 2020–following the pandemic and introduction of population-wide telehealth–compared to 2019, particularly for medically underserved groups or those known to be at risk of severe disease.

## METHODS

### Data sources and study population

We used data from the Multi-Agency Data Integration Project (MADIP), a secure data asset combining information on health, education, government payments, personal income tax, and population demographics (including the Census). Underpinning MADIP data is the Person Linkage Spine, used to create a person-level identification key by linking data from three administrative databases— Medicare Consumer Directory (records of those covered by Medicare), Department of Social Services Data and the Personal Income Tax database—resulting in virtually complete coverage of the resident population [16]. High population coverage enables high quality linkages of other datasets to the Spine. Linkage was performed by the Australian Bureau of Statistics, the Accredited Integrating Authority for this asset. For this study we used 2016 Census of Population and Housing linked to Medicare Benefits Schedule (MBS) (1 January 2019-31 December 2020) and Death Registrations (to 2019) data. Linkage was performed using deterministic and probabilistic linking methods, using name, full date of birth, address and sex, with linkage rates of 92% for Census data and 97% for deaths [16]. A direct link exists between MBS data and the Spine.

The scope of the 2016 Census was usual residents of Australia on the night of 9 August 2016 living in private and non-private dwellings [17]. It had an estimated person response rate of 94.8%, with some variation in response by ethnicity and location [18]. MBS data contain information relating to claims for medical services that are reimbursed under Medicare, including visits to GPs and other doctors outside a hospital (identified by specific MBS item numbers). Death Registrations data contain information on month and year of death for all deaths registered in Australia, and for this project were available for the 2016 to 2019 calendar years [19].

Our study population included individuals with a 2016 Census record that linked to the Spine and who had at least one MBS service claim in 2019/2020, excluding those who had a death recorded (before end of 2019, latest available death data).

### Variables

We derived the following outcome variables, based on MBS services claimed in 2019 and 2020: any use of GP services and any use of telehealth services in Q2-4 of 2020 (i.e. following introduction of telehealth items) and in Q2-4 of 2019; number of face-to-face, telehealth and total GP services by quarter for 2019 and 2020; and out-of-pocket cost (OPC) per face-to-face, telehealth and total GP services by quarter for 2019 and 2020 (see supplementary files for included MBS item numbers). We used Census data to measure characteristics of the study population), including characteristics that were markers of being medically at high-risk of poor COVID-related outcomes and other priority groups for telehealth initiatives (see Table 1 for details).

**Table 1.** Characteristics of study population, number and percentage in each sub-population.

| Characteristic | n | % |
| --- | --- | --- |
| **Total** | 19116734 | 100 |
| **Age at Jan 1 2019** |  |  |
| 3-14y | 3002936 | 15.7 |
| 15-24y | 2305386 | 12.1 |
| 25-44y | 5141525 | 26.9 |
| 45-69y | 6210584 | 32.5 |
| ≥70y | 2456303 | 12.8 |
| **Sex** |  |  |
| Male | 9163494 | 47.9 |
| Female | 9953240 | 52.1 |
| **Education** |  |  |
| University degree | 3423537 | 26.0 |
| Yr12 or dip/cert | 6080518 | 46.1 |
| No Yr12 or dip/cert | 3369575 | 25.6 |
| **Income** |  |  |
| \$104k+ | 1785626 | 9.3 |
| \$65k-<\$104k | 3622996 | 19.0 |
| \$26k-<\$65k | 7899299 | 41.3 |
| \$1 -<\$26k) | 3330954 | 17.4 |
| **Region** |  |  |
| Metro | 13687690 | 71.6 |
| Inner Regional | 3481223 | 18.2 |
| Outer regional/Remote | 1887422 | 9.9 |
| **Marital status** |  |  |
| Partnered | 9113613 | 61.6 |
| Single | 5675997 | 38.4 |
| **English Proficiency** |  |  |
| Proficient | 14174751 | 95.8 |
| Not proficient | 464316 | 3.1 |
| **Employment status** |  |  |
| Employed | 9245254 | 62.5 |
| Unemployed/ underemployed | 5360630 | 36.3 |

Notes: Yr12, year 12 of high school, dip/cert, diploma or certificate. For education, only those 25 years or older at Census are included in the analysis (n=13179119), for marital status, English proficiency and employment status only those 18 years or older at Census are included in the analysis (n=14789610). Missing/not applicable; income 12.96% [includes response not applicable, nil reported, partial income reported], education 2.32%, region 0.32%, English proficiency 1.02%, employment status 1.24%.

### Analysis

We calculated: proportion of people using services in Q2-4 of 2019 and in Q2-4 of 2020; mean number of services per person per quarter in 2019 and 2020, and mean OPC per service per quarter in 2019 and 2020. We did this for the total sample, and separately by population subgroup, using regression models to adjust for age (in 10-year groups) and sex, logistic regression to estimate proportion using services, zero-inflated negative binomial regression to estimate mean number of services, and generalised linear models with a gamma distribution and logit link function to estimate mean OPC per service. To describe changes over time, we calculated and plotted the adjusted mean number of services and the ratio of the adjusted mean number of services in 2020 vs 2019 by quarter, for total GP services and separately for face-to-face and telehealth services. We did the same for OPC per service. We also plotted the proportion of GP services by telehealth by quarter in 2020.

In supplementary analyses, we repeated the analyses separately for the state of Victoria (given greater community transmission of the virus in this state and more stringent public health measures compared to the remainder of Australia) and the remainder of Australia.

In sensitivity analyses we repeated the analyses to include those who died as these participants may differ in their sociodemographic and health risk profile.

Stata version 15.1 was used for all analyses, completed in DataLab, a secure remote access computer facility for analysis of data compiled and managed by the ABS.

We obtained ethics approval for this study from The Australian National University Human Research Ethics Committee (HREC 2020/577).

## RESULTS

### Sample characteristics

After excluding Census participants whose data did not link to the Spine (n=2,650,356), who died or had invalid death dates (n=279,654), or did not have an MBS service in 2019 or 2020 (n=1,636,280), the final study population included 19,147,620 individuals (∼81% of the Census population, Table 1).

### Any use of GP services

Overall, 86% of people saw a GP in Q2-4 of 2020, compared to 90% in the same period in 2019. In both periods, these proportions varied across population subgroups, consistent with that typically found for health service use, with service use higher among females, older ages and more disadvantaged groups. (Table 2).

**Table 2.** Proportion of persons with any use of GP MBS services in Q2-Q4 of 2019 and 2020, total and for telehealth, by sociodemographics.

|  | Proportion (%) using GP services (95%CI) |  |  | Proportion (%) using at least one GP telehealth service (95%CI) |
| --- | --- | --- | --- | --- |
|  | Q2-Q4 2019 | Q2-Q4 2020 | Change | Q2-Q4 2020 |
| **Total** | 89.5(89.5-89.5) | 85.5(85.4-85.5) | -4.0 | 48.7(48.6-48.7) |
| **Characteristic** |  |  |  |  |
| **Age at Jan 1 2019** |  |  |  |  |
| 3-14y | 85.3(85.2-85.3) | 75.9(75.9-76.0) | -9.4 | 30.8(30.8-30.9) |
| 15-24y | 85.2(85.2-85.3) | 80.7(80.7-80.8) | -4.5 | 41.8(41.7-41.9) |
| 25-44y | 87.8(87.8-87.8) | 84.1(84.1-84.2) | -3.7 | 49.0(48.9-49.0) |
| 45-69y | 91.5(91.5-91.6) | 90.0(90.0-90.0) | -1.5 | 53.8(53.8-53.8) |
| ≥70y | 96.8(96.8-96.8) | 92.6(92.6-92.6) | -4.2 | 62.4(62.4-62.5) |
| **Sex** |  |  |  |  |
| Male | 86.8(86.8-86.8) | 82.3(82.2-82.3) | -4.5 | 42.6(42.6-42.6) |
| Female | 92.0(92.0-92.0) | 88.4(88.4-88.5) | -3.6 | 54.3(54.2-54.3) |
| **Education** |  |  |  |  |
| University degree | 90.6(90.6-90.7) | 87.6(87.6-87.7) | -3.0 | 54.0(53.9-54.0) |
| Yr12 or dip/cert | 91.5(91.5-91.5) | 89.0(89.0-89.0) | -2.5 | 54.2(54.2-54.3) |
| No Yr12 or dip/cert | 92.0(91.9-92.0) | 89.4(89.4-89.4) | -2.6 | 53.6(53.6-53.7) |
| **Income** |  |  |  |  |
| \$104k+ | 88.6(88.5-88.6) | 84.6(84.6-84.7) | -4.0 | 48.1(48.1-48.2) |
| \$65k-<\$104k | 89.6(89.6-89.6) | 85.6(85.6-85.6) | -4.0 | 49.1(49.1-49.2) |
| \$26k-<\$65k | 89.7(89.6-89.7) | 85.7(85.7-85.7) | -4.0 | 48.9(48.9-49.0) |
| \$1 -<\$26k | 90.0(89.9-90.0) | 85.9(85.8-85.9) | -4.1 | 48.6(48.6-48.7) |
| **Region** |  |  |  |  |
| Metro | 90.1(90.1-90.2) | 85.9(85.9-85.9) | -4.2 | 49.6(49.5-49.6) |
| Inner Regional | 88.3(88.3-88.3) | 84.8(84.8-84.9) | -3.5 | 49.4(49.4-49.5) |
| Outer regional/Remote | 86.8(86.8-86.9) | 83.5(83.4-83.5) | -3.3 | 40.9(40.8-40.9) |
| **Marital status** |  |  |  |  |
| Partnered | 91.1(91.1-91.1) | 88.5(88.5-88.5) | -2.6 | 53.5(53.4-53.5) |
| Single | 90.5(90.5-90.5) | 87.2(87.1-87.2) | -3.3 | 52.7(52.7-52.8) |
| **English Proficiency** |  |  |  |  |
| Proficient | 90.8(90.8-90.9) | 88.0(88.0-88.0) | -2.8 | 53.6(53.6-53.7) |
| Not proficient | 92.0(91.9-92.1) | 87.5(87.4-87.6) | -4.5 | 40.5(40.4-40.6) |
| **Employment status** |  |  |  |  |
| Employed | 90.6(90.5-90.6) | 87.7(87.7-87.8) | -2.8 | 52.8(52.8-52.8) |
| Unemployed/underemployed | 91.7(91.7-91.8) | 88.5(88.5-88.6) | -3.2 | 54.0(54.0-54.1) |

Notes: Any use indicates a person at least one MBS service of that type during that period. Yr12, year 12 of high school, dip/cert, diploma or certificate. For education only those 25 years or older at Census are included in the analysis (n=13179119), for marital status, English proficiency and employment status only those 18 years or older at Census are included in analysis (n=14789610). Predicted estimated proportions are adjusted for age (in 10-year groups) and sex.

Almost half of the population (49%) used GP telehealth services at least once in Q2-4 of 2020, up from < 2% in the same period in 2019. Generally, groups who used more GP services (e.g. people aged ≥70, those with lower household incomes) were also more likely to use telehealth at least once, with one exception. Patients not proficient in English were more likely to use GP services than patients proficient in English (with similar proportions using GP services in 2020), but were less likely to be users of telehealth (Table 2).

### Number of GP services

The mean number of face-to-face services per person per quarter in 2020 declined after Q1 (start of pandemic and introduction of widescale telehealth), while the mean number of telehealth services per patient increased, with total GP services per patient similar across quarters. This pattern was generally consistent across all sociodemographic groups examined (supplementary file).

Across most sociodemographic groups, individuals used more GP services in 2020 compared to 2019, i.e. ratio of services in 2020:2019>1, with ratios generally highest in Q3 and Q4 (Figure 1). However, individuals aged 3-14 and ≥70 years and those with limited English proficiency used fewer GP services in 2020 than 2019.

**Figure 1:**
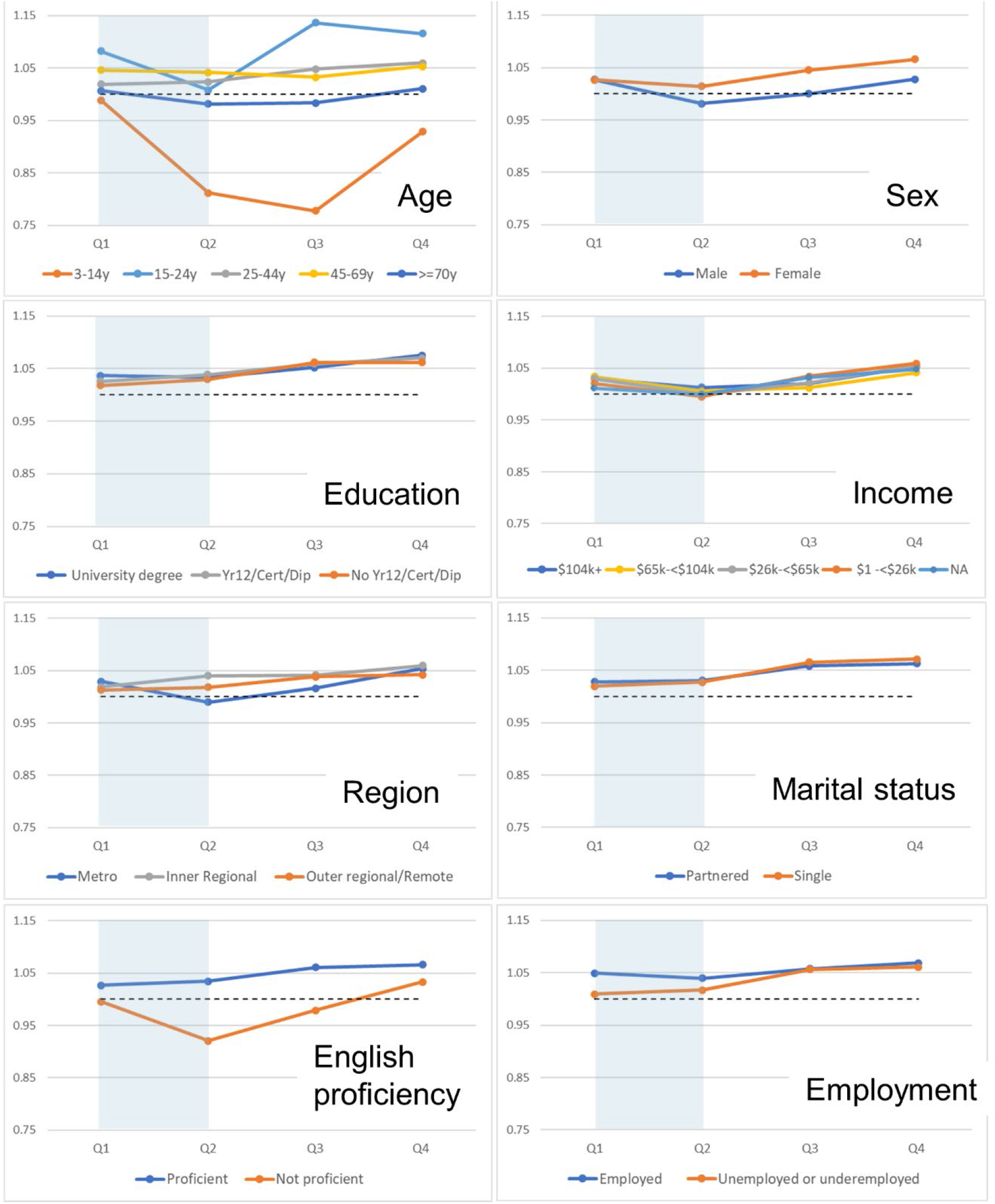
Ratio of mean number of MBS GP services per person 2020 vs 2019, by quarter and sociodemographics. Notes: Abbrev. GP, general practitioner; MBS, Medicare Benefits Schedule; Yr12, year 12 of high school; dip/cert, diploma or certificate. For education only those 25 years or older at Census are included in analysis (n=13179119), for marital status, English proficiency and employment status only those 18 years or older at Census are included in analysis (n=14789610). Ratio of 2020 to 2019 predicted estimated mean number of services adjusted for age (10-year groups) and sex. Dashed line represents a ratio of 1 where the mean estimate in 2020 was the same as 2019. Telehealth initiative introduced during March 2020 (shaded area).

Across all groups, the proportion of total services by telehealth peaked at 30-35% before stabilising at ∼25%, but was lower among older people, males, and those with low education, low income, living in outer regional/remote areas and with limited English proficiency (Figure 2).

**Figure 2.**
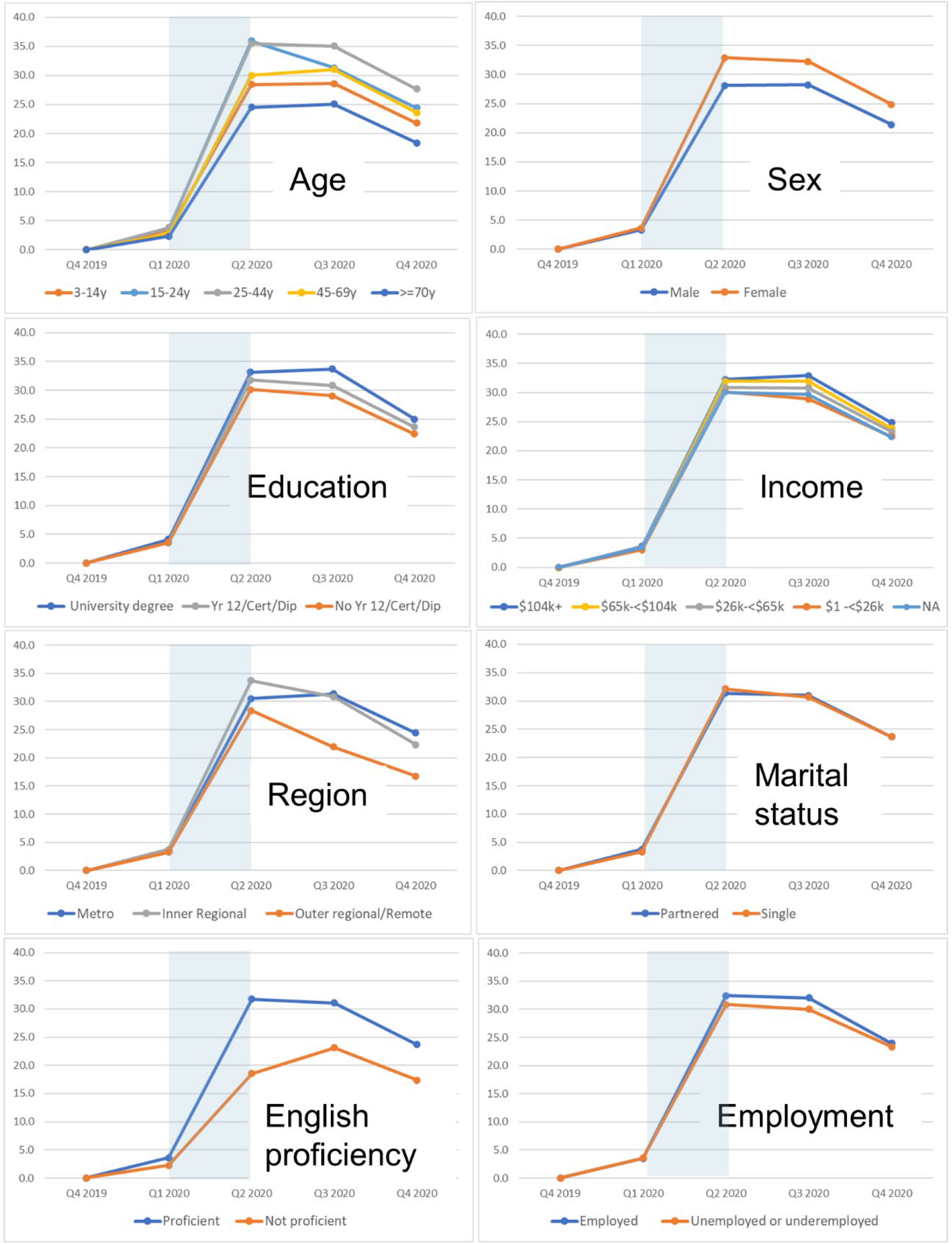
Proportion of total GP services per person by telehealth, by quarter and sociodemographics. Notes: Abbrev. GP, general practitioner; MBS, Medicare Benefits Schedule; Yr12, year 12 of high school; dip/cert, diploma or certificate. For education only those 25 years or older at Census are included in analysis (n=13179119), for marital status, English proficiency and employment status only those 18 years or older at Census are included in analysis (n=14789610). Proportion of predicted estimated mean number of services by telehealth, adjusted for age (10-year groups) and sex, by quarter. Telehealth initiative introduced during March 2020 (shaded area).

### Out-of-pocket costs

The mean OPC per service in 2020, for face-to-face services and total GP services in 2020 declined in Q2, from $5 AUD in Q1 to $2.60 AUD in Q2, before increasing in Q3 and Q4 but remaining below those in Q1 (supplementary file). Across all sociodemographic groups, mean OPC per service was lower in Q2-Q4 of 2020 compared to 2019, i.e. ratio of OPC in 2020:2019<1, with ratios lowest in Q2 (Figure 3). The largest decreases in OPC per service were found for individuals aged 3-14 and ≥70 years, residing in major cities and with limited English proficiency. For telehealth services specifically, mean OPC per service was very low ($0.15 AUD in Q1 2020), but increased over the latter part of 2020, across all groups.

**Figure 3.**
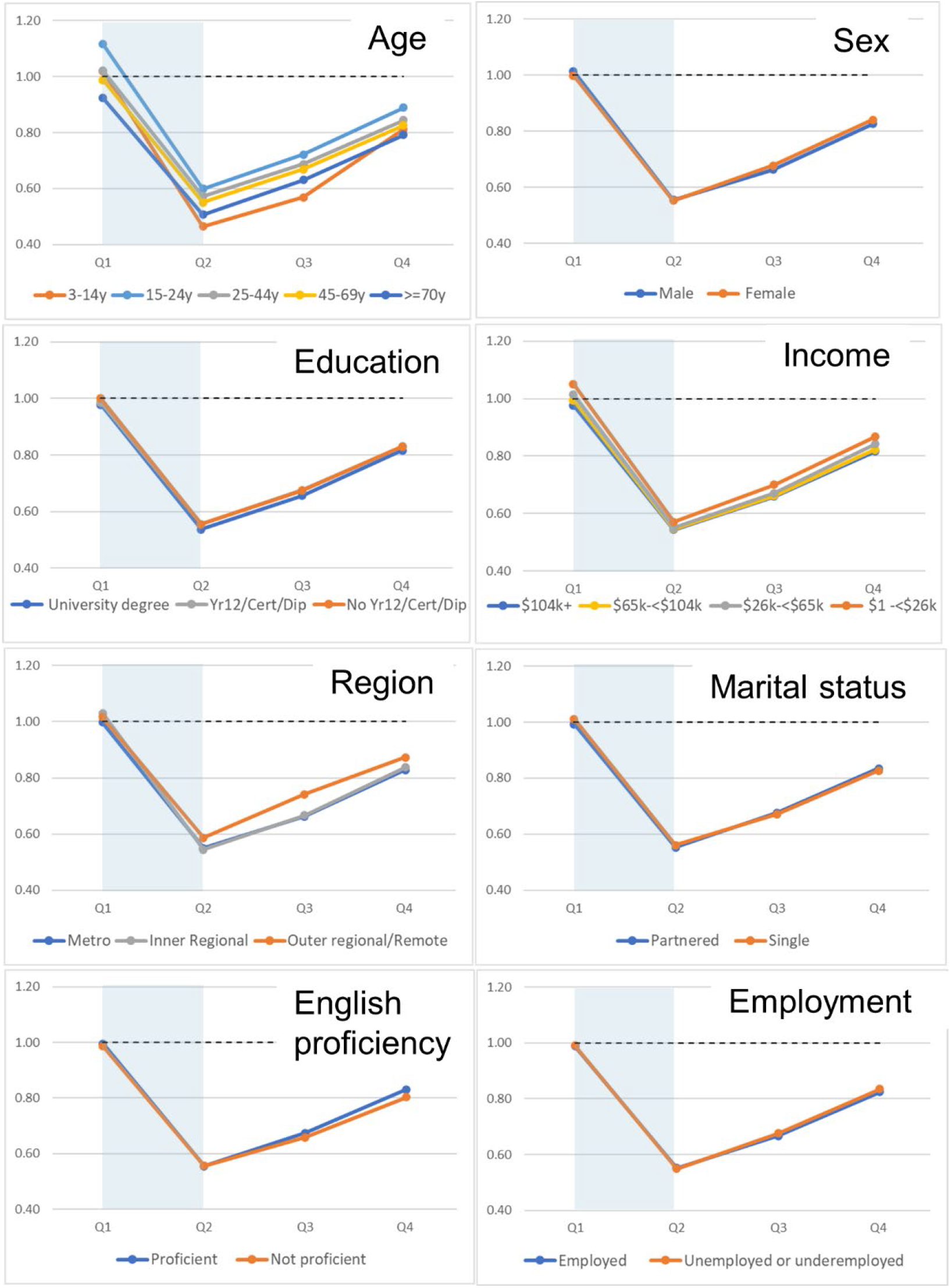
Ratio of mean OOP cost per service 2020 vs 2019, by quarter and sociodemographics. Notes: Abbrev. GP, general practitioner; MBS, Medicare Benefits Schedule; Yr12, year 12 of high school; dip/cert, diploma or certificate. For education only those 25 years or older at Census are included in analysis (n=13179119), for marital status, English proficiency and employment status only those 18 years or older at Census are included in analysis (n=14789610). Ratio of 2020 to 2019 predicted estimated mean out-of-pocket cost per service adjusted for age (10-year groups) and sex. Dashed line represents a ratio of 1 where the mean estimate in 2020 was the same as 2019. Telehealth initiative introduced during March 2020 (shaded area).

In supplementary and sensitivity analyses, patterns of use and costs were similar to the main analysis (supplementary files). However, when analysed separately for Victoria, the proportion of total services by telehealth peaked at ∼50% in Q2 2020, while OPC did not rise again until Q4 2020.

## DISCUSSION

### Summary of findings

In the first year of the pandemic in Australia following the introduction of new telehealth items, use of GP services was generally maintained while OPC were minimised. However, this was not the case for all groups; total GP use was lower for 3-14 year olds, older people, and people with limited English proficiency. Around one-quarter of all services were delivered by telehealth, and those who typically used more health services were also more likely to use telehealth, excepting those with low English proficiency. However, some key groups, often disadvantaged or medically underserved populations, had a lower proportion of services delivered via telehealth, including older people, males, people with low education or low income, those living in regional/remote areas and those with low English proficiency. OPC per service dropped substantially in the early months of the pandemic, with limited recovery by the end of 2020. While this may have been crucial in supporting patient access to care, it presents a potential risk for the ongoing viability of general practice.

### Comparison with existing literature

Previous Australian studies have shown that overall levels of GP services were maintained in the early months of the pandemic, largely through substitution of in-person services with telehealth [5, 6, 20]; none examined whether this varied across population subgroups. In other countries with rapidly increased access to population-wide telehealth, levels of ambulatory care were substantially reduced[7-10, 14]. Throughout 2020, Australia was still attempting eradication of the COVID-19 virus and case numbers were comparatively low[1], which may explain these differences. Patterns of telehealth uptake by subpopulation are similar to that reported for from Australian primary care clinic data[13] and the US[7, 10-12].

The decrease in use and uptake of telehealth found in our study for some users may be due to policy, technological, social or other barriers or due to pandemic-related changes in healthcare use. For example, patient preference for virtual care when their provider knows them well[21], or confidence in using virtual care technology[22]. Further, a US study found that practice and clinician factors accounted for more of the variation in video use (versus audio-only) than patient-level factors [23]. Providers report a lack of technology or the need for physical examination as reasons for preferring in-person consultations[8, 24], while the background risk of COVID[8, 21, 24] and public health directives[8] influence both provider and patient preferences for in-person vs virtual consultations. Further work is needed to better understand the interplay of patient, provider/practice and system-level factors and whether access is equitable and according to need.

### Strengths and limitations

This study used whole-of-population, administrative data to report on use and costs of GP services during the first three quarters of the COVID-19 pandemic. Given the direct link between the MADIP Spine and MBS data, we expect complete ascertainment of all outcomes among the study population. Indicators of health care need, including health conditions, were not available in the data. For this reason, we relied on markers (e.g. age) to indicate those medically at-risk of poor outcomes and those from traditionally underserved populations. Census data were collected in August 2016; as such, a degree of change in time varying characteristics is likely resulting in misclassification. Furthermore, given data availability, we could not fully account for deaths to exclude those from the study population who may have been out of scope. This may have impacted results among older people, potentially explaining (at least to some degree) some of the reduction in use of GP services in 2020.

### Policy implications and future research

The findings suggest that the rapid transition in primary healthcare to whole-of-population telehealth has been effective in supporting access to care for the majority of the Australian population. Arguably, need for care may have been greater in the first year of the pandemic as COVID spread, in addition to the increase in mental health conditions related to the impact of disease control policies (such as social isolation, financial insecurity). Nevertheless, the current findings highlight the risk that some groups, specifically those with generally greater health needs but who typically find services harder to reach, may be missing out. Further work is required to understand and overcome potential barriers to use of telehealth, particularly for children and older people, and individuals with low English proficiency. Moreover, while most of the population were able to access services, the impact on quality-of-care and safety is unknown.

The reduction in the cost of services for all groups likely offset a significant barrier to care due to widespread economic disruption experienced during the pandemic. While these current findings indicate OPC started to increase, this had not yet reached pre-pandemic pre-telehealth levels. Given the limited investment in general practice in the last decade[25, 26], ongoing integration of telehealth will need to balance affordability of services with the sustainability of general practices.

### Conclusion

This study identified subpopulations, including those who are medically at-risk or underserved, with lower uptake of telehealth services in the first year of the pandemic and can be used to assist GPs and local community health services to tailor how they integrate telehealth with usual clinical practice. It can also inform policy responses required to ensure telehealth achieves the anticipated objectives. Our findings regarding OPC raise potential issues related to the ongoing sustainability and capacity of general practice to continue to meet the healthcare needs of Australians, and the need to account for this with future widescale integration of telehealth.

## Data Availability

Data part of the Multi-Agency Data Integration Project are available for approved projects to approved government and non-government users. https://www.abs.gov.au/websitedbs/D3310114.nsf/home/Statistical+Data+Integration+-+MADIP

## Additional Information

### Ethics approval and consent to participate

Ethics approval for this project was obtained from the Australian National University Human Research Ethics Committee (2020/577).

### Authors’ contributions

DB, KD, GJ, RK conceived and designed the analysis. DB completed data analysis and drafted the manuscript. MB contributed to data analysis. JW and AD contributed to interpretation and drafting of the manuscript. All authors revised the work for intellectual content and approved the final version of the manuscript.

## Acknowledgements

We acknowledge the contributions of members the ANU Telehealth in Primary Care group: Christine Phillips, Emily Banks, Jason Agostino, Emily Lancsar, Jane Desborough, Sally Hall-Dykgraaf, Anne Parkinson, Kay Soga, Dan Chateau, Nina Lazaveric, and Hsei-Di Law.

